# Immunogenicity of COVID-19 mRNA Vaccines in Patients with Acute Myeloid Leukemia and Myelodysplastic Syndrome

**DOI:** 10.1101/2022.01.26.22269932

**Authors:** David C. Helfgott, Sabrina Racine-Brzostek, Kelsey J. Short, Zhen Zhao, Paul Christos, Itzel Nino, Tina Niu, Jorge Contreras, Ellen K. Ritchie, Pinkal Desai, Michael Samuel, Gail J. Roboz

## Abstract

Immunocompromised patients are particularly susceptible to serious complications from infection with severe acute respiratory syndrome coronavirus-2 (SARS-CoV-2). Two mRNA vaccines, BNT162b2 and mRNA-1273, have been shown to have excellent clinical efficacy in immunocompetent adults, but diminished activity in immunocompromised patients. In this study, we measured anti-spike SARS-CoV-2 antibody response, avidity, and surrogate neutralizing antibody activity in Coronavirus Disease 2019 (COVID-19) vaccinated patients with acute myeloid leukemia (AML) and myelodysplastic syndrome (MDS). Anti-spike SARS-CoV-2 antibody was present in 89% of AML and 88% of MDS patients, but median antibody levels for were lower than in healthy controls (p=0.001 and p=0.04, respectively). SARS-CoV-2 antibody avidity and neutralizing antibody activity from AML patients were significantly lower than controls (p=0.028 and p=0.002, respectively). There was a trend toward higher anti-spike SARS-CoV-2 antibody levels after mRNA-1273 vaccination. Antibody avidity was greater in patients after mRNA-1273 versus BNT162b2 (p=0.01) and there was a trend toward greater neutralizing antibody activity after mRNA-1273 versus BNT162b2 vaccination.

## Introduction

Immunocompromised patients are particularly susceptible to serious complications from infection with severe acute respiratory syndrome coronavirus-2 (SARS-CoV-2) (1). In December 2020 two mRNA vaccines became available, promising to change the trajectory of Coronavirus Disease 2019 (COVID-19). The BNT162b2 (BioNTech/Pfizer) and mRNA-1273 (Moderna) vaccines were demonstrated in clinical trials to have clinical efficacies of 95% (2) and 94% (3) respectively in immunocompetent adults. This protection was replicated in “real-world” experience as people began to receive vaccinations (4).

Immunocompromised patients may have suboptimal antibody responses to vaccinations (5). Evidence has begun to accumulate that certain immunocompromised populations have an impaired antibody response to COVID-19 vaccination. Haberman et al showed that patients with immune-mediated inflammatory diseases receiving treatment with methotrexate developed lower levels of anti-spike SARS-CoV-2 IgG in response to BNT162b2 vaccine compared to both healthy controls and similar patients who did not receive methotrexate (6). Deepak et al found that patients with chronic inflammatory diseases treated with glucocorticoids were more likely not to seroconvert or had a much lower anti-spike SARS-CoV-2 IgG titer in response to mRNA vaccines compared to healthy individuals; they also found lower antibody titers in patients receiving Janus kinase inhibitors and in patients receiving rituximab or ocrelizumab (7). Several studies have demonstrated that solid organ transplant recipients receiving anti-rejection therapy including cyclosporine, steroids, mycophenolate, and calcineurin blockers developed very low levels of anti-spike SARS-CoV-2 antibodies after vaccination, or more commonly had a negative antibody response to COVID-19 mRNA vaccines (8,9,10,11).

Patients with hematologic malignancies may have disease-specific immune defects which could impair antibody production. Patients with active multiple myeloma who received BNT162b2 vaccine were less likely to seroconvert and those that did had lower anti-spike SARS-CoV-2 antibody titers compared to patients with smoldering myeloma or healthy controls (12). Patients with chronic lymphocytic leukemia who received BNT162b2 vaccine had a very low seroconversion rate and, for those who did seroconvert, the median anti-spike SARS-CoV-2 antibody titer was about 15% that of healthy controls (13). In that study, no patient with CLL who received anti-CD20 treatment in the 12 months before vaccination developed antibodies (13). In patients with non-Hodgkin lymphomas, Perry et al. found a very low seroconversion rate and very low antibody titers in patients receiving anti-CD20 therapy and significantly lower antibody titers in untreated or previously anti-CD20 treated patients, compared to healthy controls (14). Greenberger et al. found that, while patients with Hodgkin’s lymphoma all seroconverted after COVID-19 vaccination, the seroconversion rate in patients with non-Hodgkin subtypes was very low (15). This group found a high seroconversion rate in patients with myeloma, chronic myelocytic leukemia, acute lymphocytic leukemia, and acute myeloid leukemia (15).

Effective antibody response to vaccination likely involves more than just measurable antibody level. Antibody avidity (functional affinity), which is the total strength of the interactions between antibody and antigen, and neutralizing antibody titer, which reflects the ability of an antibody to prevent an antigen from interacting with its target, are also likely important determinants of vaccine efficacy. SARS-CoV-2 antibody avidity has been measured after natural infection (16,17,18), but there are no avidity studies reported in vaccinated patients with hematologic malignancies. Neutralizing antibody activity has been studied in vaccinated patients with hematologic malignancies. Vaccinated patients with solid tumors have exhibited higher SARS-CoV-2 neutralizing antibody titers compared to vaccinated patients with hematologic malignancies (19,20). Patients with multiple myeloma (21) and Waldenstrom’s macroglobulinemia (22) had less SARS-CoV-2 neutralizing antibody activity compared to controls on days 22 and 50 after BNT162b2 vaccination. Chung et al. found that at both 1 month and 3 months after vaccination with an mRNA vaccine, patients with hematologic malignancies (almost all CLL, lymphoma, and myeloma) had lower neutralizing antibody activity compared to controls (23).

In this study, we measured the anti-spike SARS-CoV-2 antibody response, antibody avidity, and surrogate neutralizing antibody activity in COVID-19 vaccinated patients with acute myeloid leukemia (AML) and myelodysplastic syndrome (MDS).

### Patients and Methods

The study was approved by the Weill Cornell Medicine Institutional Review Board (IRB# 21-02023288, 20-11022929). Serum samples were collected from adult outpatients with AML and MDS who received two doses of an mRNA Covid-19 vaccine at least 14 days prior to serum collection and who had no history of a positive SARS-CoV-2 polymerase chain reaction (PCR) test. Patient age, underlying disease, medication history, COVID-19 vaccine history, and absolute neutrophil and lymphocyte counts closest to the dates of vaccination were recorded. Serum samples were stored at -20^°^C until assayed.

Control samples were collected from health care workers at the same institution without prior evidence of COVID-19 infection who completed both doses of BNT-162b2 or mRNA-1273 vaccine. Eighteen healthy health care workers who received BNT162b2 vaccine were matched for both age and time from completion of vaccination to serum collection to the eighteen study patients who seroconverted after BNT162b2 vaccination. Of the control health care workers, only 11 received mRNA-1273 vaccine and these are all included.

Antibodies to SARS-CoV-2 spike (S) receptor binding domain (RBD) antigen and SARS-CoV-2 nucleocapsid (N) antigen were detected using the Elecsys Anti-SARS-CoV-2S and Anti-SARS-CoV-2N electrochemiluminescence immunoassays performed on the Roche Cobas e411 (Roche Diagnostics, Indianapolis IN).

Avidity and surrogate neutralizing antibody titers were measured as previously described (24,25). The avidity assay measures the relative dissociation rate (dR) of SARS-CoV-2 antibodies from the SARS-CoV-2 spike RBD, which is inversely related to antibody avidity. The surrogate neutralizing antibody (sNAb) assay is a competitive binding assay based on SARS-CoV-2 antibody-mediated inhibition of the interaction between the angiotensin-converting enzyme 2 (ACE2) protein and the RBD; the assay result is the percentage of RBD-ACE2 binding (therefore the lower the %RBD-ACE2, the higher the SARS-CoV-2 neutralizing activity). This sNAb assay has been shown to correlate with the SARS-CoV-2 plaque reduction neutralization test and the pseudo virus neutralization test (25).

Statistical analysis was performed using GraphPad Prism version 9. The Mann-Whitney U test and Kruksal-Wallis test were used for group comparisons of interest (i.e., MDS, AML, MDS+AML, healthy controls). All p-values are two-sided with statistical significance evaluated at the 0.05 alpha level. Ninety-five percent confidence intervals (95% CI) were calculated for all parameters of interest to assess the precision of the obtained estimates. Pairwise group comparisons were not corrected for multiple comparisons due to the pilot (hypothesis-generating) nature of the study.

## Results

Serum samples were collected from 24 AML and 21 MDS patients. Anti-nucleocapsid antibody was positive in 6 patients (2 AML and 4 MDS). These patients were excluded, and 22 AML and 17 MDS patients were included in the final analysis. The median time from completion of vaccination to sample collection was 37 days (range 14-102 days) for the patients with AML and 48 days (range 14-82 days) for those with MDS. The median age was 74.5 years (range 34-88 years) for AML patients and 78 years (range 66-91 years) for MDS patients. Baseline patient characteristics are shown in Table 1.

**Table 1.** Patient Characteristics

|  | All Patients (n=39) | AML (n=22) | MDS (n=17) |
| --- | --- | --- | --- |
| **Age at Diagnosis (years)** |  |  |  |
| Median | 76 | 74.5 | 78 |
| Range | 34-91 | 34-88 | 66-91 |
| **Gender** |  |  |  |
| Female | 20 (51%) | 14 (64%) | 6 (35%) |
| Male | 19 (49%) | 8 (36%) | 11 (65%) |
| **Time from second vaccination to sample (days)** |  |  |  |
| Median | 45 | 37 | 48 |
| Range | 14-102 | 14-102 | 14-82 |
| **ELN Risk (AML)<sup>1</sup>** |  |  |  |
| Favorable |  | 5 (24%) |  |
| Intermediate |  | 6 (28%) |  |
| Adverse |  | 10 (48%) |  |
| **IPSS-R (MDS)<sup>2</sup>** |  |  |  |
| Median |  |  | 3.14 |
| Range |  |  | 1.05-7.96 |
| Very Low |  |  | 2 (13%) |
| Low |  |  | 4 (27%) |
| Intermediate |  |  | 6 (40%) |
| High |  |  | 1 (7%) |
| Very High |  |  | 2 (13%) |
| **Disease status at first vaccination (AML)** |  |  |  |
| Remission |  | 14 (64%) |  |
| Active Disease |  | 8 (36%) |  |
| **Chemo/targeted therapy within 8 weeks before first vaccination** |  |  |  |
| Yes | 16 (41%) | 13 (59%) | 3 (18%) |
| No | 23 (59%) | 9 (41%) | 14 (82%) |
| **ANC<sup>3</sup> at first vaccination (x 10<sup>3</sup>/μL)** |  |  |  |
| Median | 1.8 | 1.9 | 1.4 |
| Range | 0.2-20.4 | 0.3-20.4 | 0.2-4.4 |
| **ALC<sup>4</sup> at first vaccination (x 10<sup>3</sup>/μL)** |  |  |  |
| Median | 0.95 | 1.0 | 0.9 |
| Range | 0.2-2.7 | 0.4-2.5 | 0.2-2.7 |
| **Covid-19 vaccination type** |  |  |  |
| BNT162b2 | 21 (54%) | 15 (68%) | 6 (35%) |
| mRNA-1273 | 18 (46%) | 7 (32%) | 11 (65%) |

<sup>1</sup> One patient with acute promyelocytic leukemia was excluded from ELN (European LeukemiaNet) Risk stratification<sup>2</sup> Two patients with chronic myelomonocytic leukemia were excluded from IPSS-R (revised international prognostic scoring system) classification<sup>3</sup> Absolute neutrophil count<sup>4</sup> Absolute lymphocyte count

All control samples were negative for anti-nucleocapsid antibody and the median time from completion of vaccination to serum collection for controls was 34 days (range 14-142 days). The median age of the healthy controls was 57 years (range 30-78 years). The median anti-spike SARS-CoV-2 antibody level for the control samples was 1192 U/ml (range 226.4-8357.0 U/ml). In the healthy controls, median avidity as measured by the relative dissociation rate of SARS-CoV-2 antibody from RBD was 8.2 × 10^−4^ /sec, and median %ACE2-RBD binding was 3.9%.

Among AML patients, 20 (91%) had measurable anti-spike SARS-CoV-2 antibody and 2 (9%) did not seroconvert (anti-spike SARS-CoV-2 antibody <0.8 U/ml). One of the two patients who did not seroconvert was receiving prednisone at the time of vaccination. The median anti-spike SARS-CoV-2 antibody level for the AML patients who seroconverted was 151.3 U/ml (range 8.99-3643.0 U/ml). In the AML patients, median avidity as measured by the dissociation rate of SARS-CoV-2 antibody from RBD was 9.4 × 10^−4^ /sec and median %ACE2-RBD binding was 25%.

Among MDS patients, 15 (88%) had measurable anti-spike SARS-CoV-2 antibody and 2 (12%) failed to seroconvert (anti-spike SARS-CoV-2 antibody <0.8 U/ml). Both patients who did not seroconvert were receiving prednisone at the time of vaccination and one of these patients had concomitant CLL and was also receiving tacrolimus and mycophenolate. The median anti-spike SARS-CoV-2 antibody level for the MDS patients who seroconverted was 250.0 U/ml (range 23.5-8474.0 U/ml). In the MDS patients, median avidity as measured by the dissociation rate of SARS-CoV-2 antibody from RBD was 8.7 × 10^−4^ /sec and median %ACE2-RBD binding was 26%.

The median anti-spike SARS-CoV-2 antibody levels for AML and MDS patients were significantly lower than those for healthy controls (figure 1). Although there was a trend toward higher antibody levels for MDS patients compared to AML patients, this was not statistically significant (figure 1). Antibody avidity was not significantly different between MDS patients and healthy controls, but SARS-CoV-2 antibodies from AML patients were significantly less avid than antibodies from the controls (p=0.028) (figure 2). SARS-CoV-2 neutralizing antibody activity was significantly lower in AML patients compared to the controls; neutralizing antibody activity in MDS patients trended lower compared to controls (figure 3). The study was underpowered to make formal AML/MDS/control comparisons of antibody level, avidity, or neutralizing antibody activity, stratified by factors such as age, time from vaccination to sample collection, remission status in AML patients, absolute neutrophil count at time of vaccinations, or absolute lymphocyte count at time of vaccinations. Treatment with decitabine (4 patients) or venetoclax (8 patients) did not appear to be related to antibody levels in AML patients, but these are small subgroups. Most patients with MDS were not receiving chemotherapy during the two months prior to the first vaccination or during the immediate post-vaccination period.

**Figure 1.**
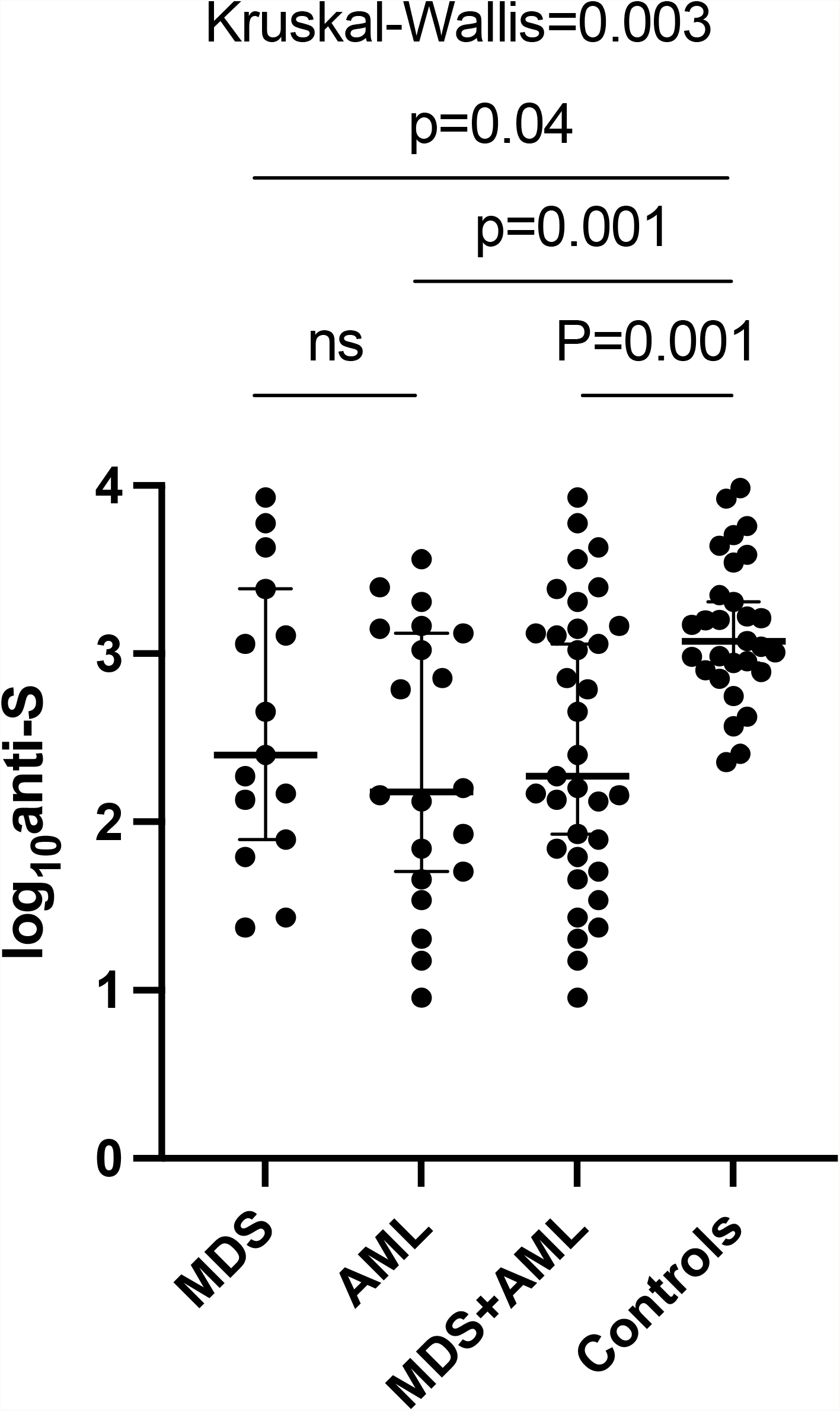
Anti-spike SARS-CoV-2 antibody levels for the patients with MDS, AML, and healthy controls, shown with median ± 95% confidence intervals. Compared to controls, the patients with MDS (p=0.04) and the patients with AML (p=0.001) had significantly lower anti-spike SARS-CoV-2 antibody levels.

**Figure 2.**
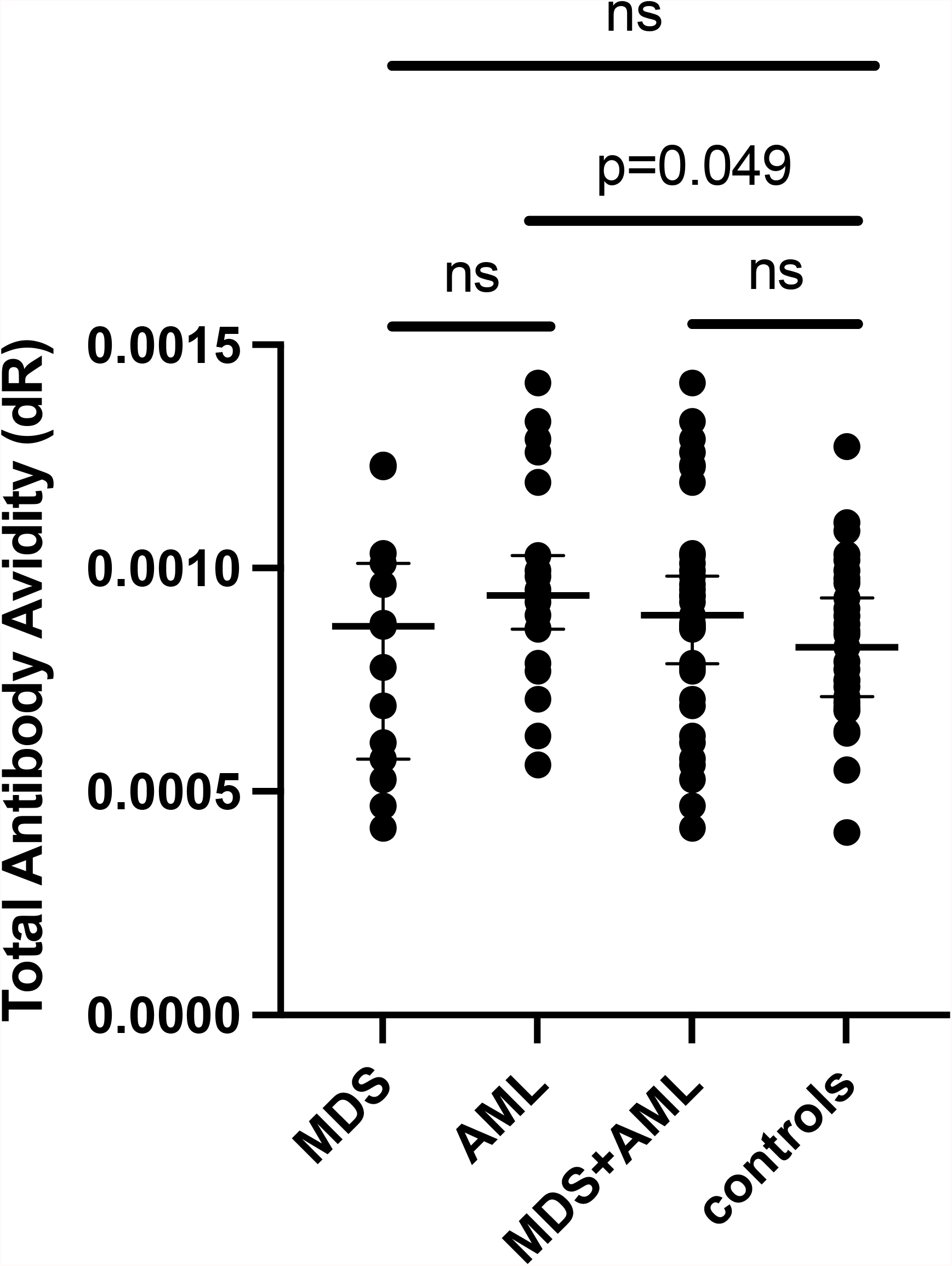
Dissociation rate of SARS-CoV-2 antibodies from RBD for the patients with MDS, AML, and healthy controls, shown with median ± 95% confidence intervals. Compared to controls, the SARS-CoV-2 antibodies from the patients with AML were significantly less avid (bound to RBD less strongly, since the dR is higher) compared to the SARS-CoV-2 antibodies from healthy controls (p=0.049).

**Figure 3.**
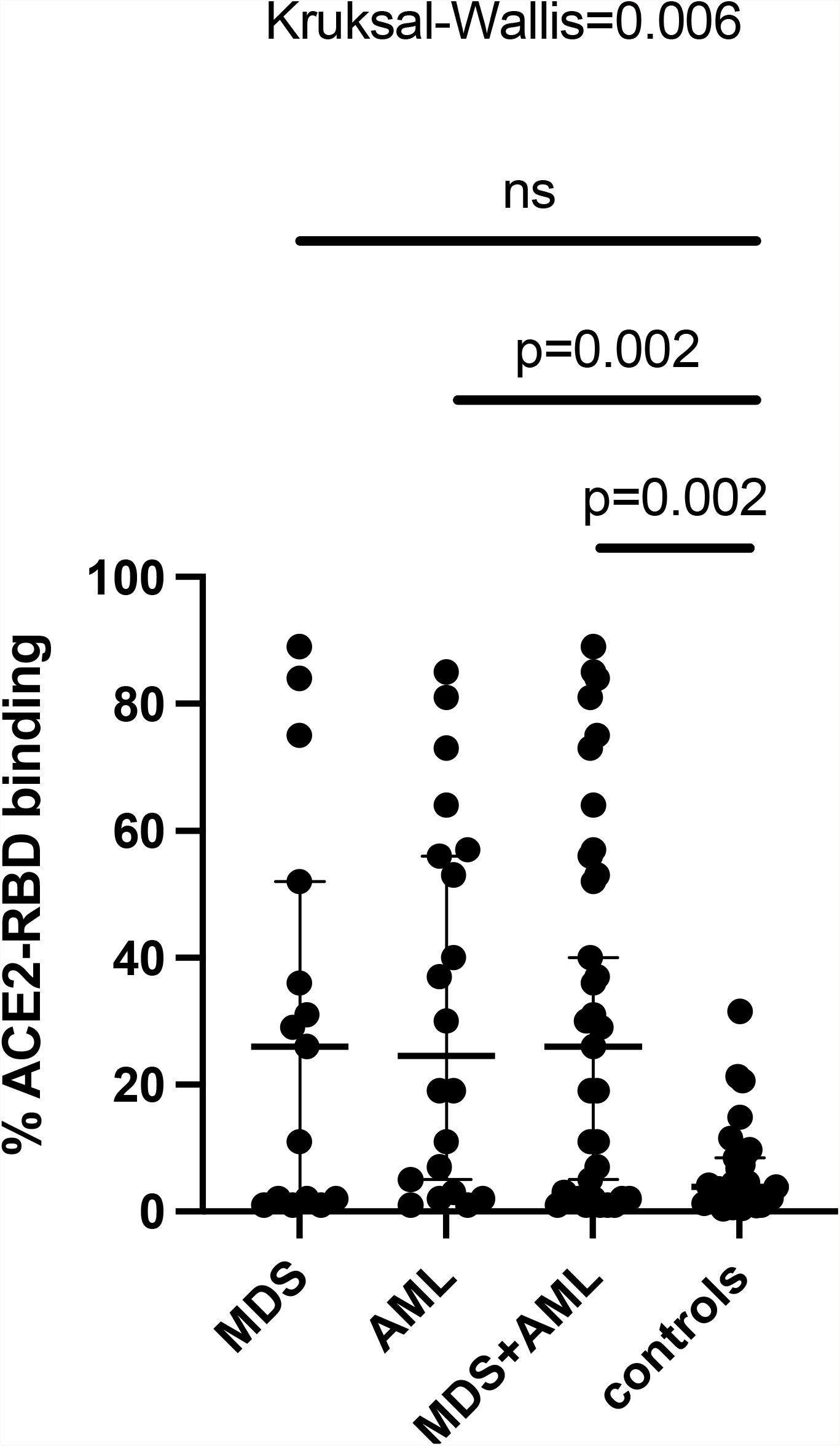
Neutralizing antibody activity as represented by the ability of SARS-CoV-2 antibodies from the patients with MDS, AML, and healthy controls to compete with the ACE2 receptor for binding with RBD, shown with median ± 95% confidence intervals. Compared to controls, the patients with AML (p=0.002) had significantly lower neutralizing antibody activity (higher %ACE-RBD binding).

There was a trend toward higher anti-spike SARS-CoV-2 antibody levels in patients who received the mRNA-1273 vaccine, but it was not statistically significant (figure 4A). For patients who received mRNA-1273 vaccine, the median anti-spike SARS-CoV-2 antibody was 452.3 U/ml and for patients who received BNT162b2 vaccine, the median anti-spike SARS-CoV-2 antibody was 108.5 U/ml. Of the 23 AML and MDS patients with anti-spike SARS-CoV-2 antibody >100 U/ml, 14 received mRNA-1273 vaccine and 9 received BNT162b2 vaccine. Among 12 patients with anti-spike SARS-CoV-2 antibody between 0.8-100 U/ml, 3 received mRNA-1273 vaccine and 9 received BNT162b2 vaccine. Antibody avidity was greater in patients who received mRNA-1273 versus BNT162b2 vaccine (figure 4B, p=0.01). Neutralizing antibody activity was greater in the patients who received mRNA-1273 versus BNT162b2 vaccine, but this was not statistically significant (figure 4C). Time from completion of vaccination to serum collection and age of seroconverted patients was similar in the two vaccine groups.

**Figure 4.**
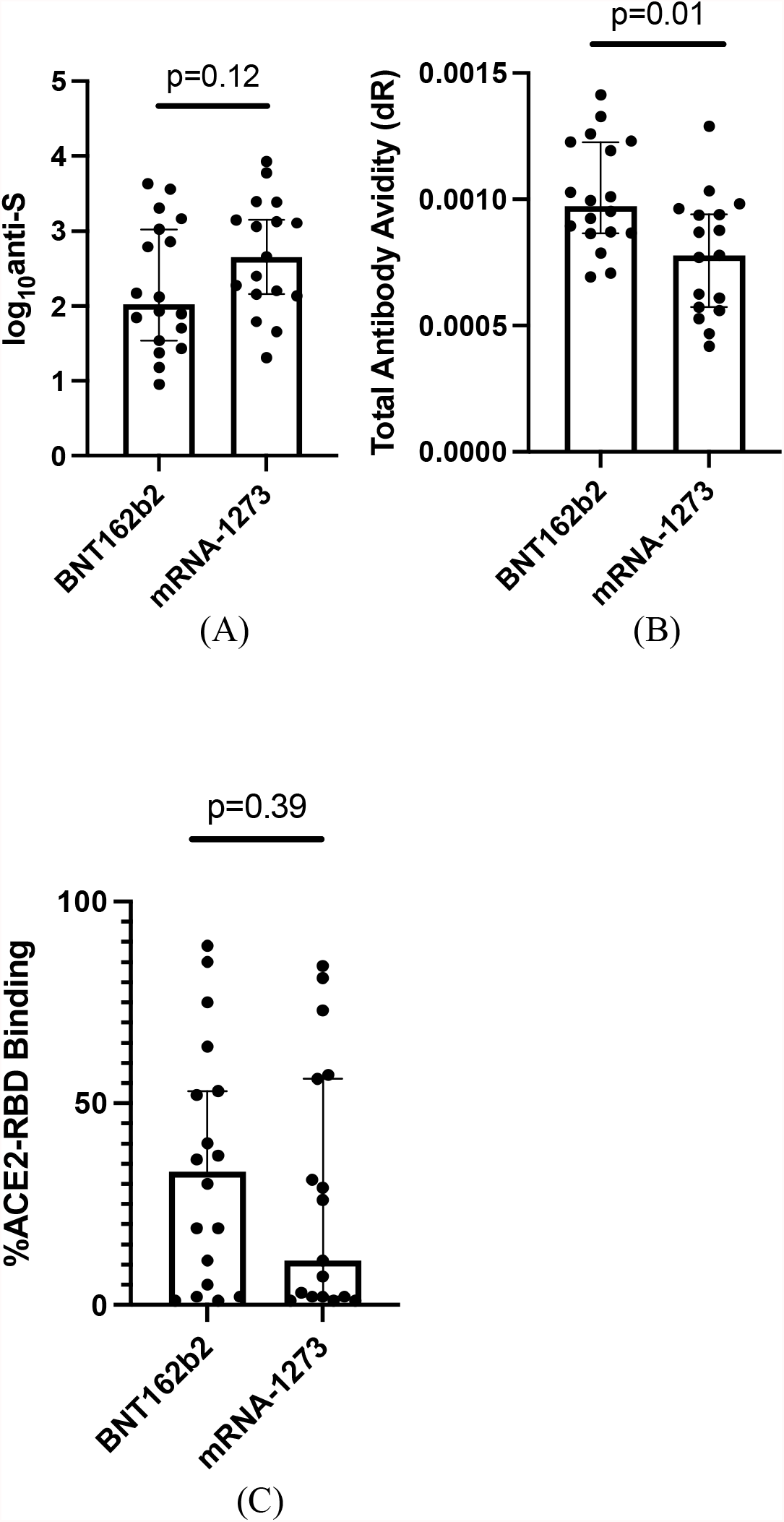
SARS-CoV-2 antibody level (A), avidity (B) and neutralizing activity (C) in the patients with who received BNT162b2 vaccine compared to the patients who received mRNA-1273 vaccine, shown with 95% confidence intervals. While there was only a trend toward higher SARS-CoV-2 antibody titer (p=0.12) and antibody neutralizing activity (p=0.39) in the patients who received mRNA-1273 vaccine, the SARS-CoV-2 antibodies from patients who received the mRNA-1273 vaccine exhibited significantly stronger binding to RBD (greater avidity) compared to the antibodies from the patients who received BNT162b2 vaccine (p=0.01).

## Discussion

In this study, 89% of patients with AML and 88% of patients with MDS developed anti-spike SARS-CoV-2 antibody at least two weeks after completing vaccination. Of the 4 non-responding patients, 3 were receiving prednisone at the time of vaccination. However, median anti-spike SARS-CoV-2 antibody titers in AML and MDS patients who seroconverted were significantly lower than in health care worker controls (figure 1). Similarly, as reported in the literature, median anti-spike SARS-CoV-2 antibody levels in healthy, uninfected controls sampled 14-21 days after mRNA vaccination and measured with the same Elecsys Anti-SARS-CoV-2 assay used in this study, are about six- and four-fold higher than the antibody levels in our AML and MDS patients, respectively. (12,13).

SARS-CoV-2 antibodies from AML patients exhibited significantly less RBD avidity than healthy controls, but there was no difference in avidity between MDS patients and controls. Of note, in this work, avidity was measured at a single time point, but in other studies of viral infections, antibody avidity has been shown to increase over time (26,27). This increase in antibody avidity, or “avidity maturation,” reflects the evolving immune response to an infection. Failure of avidity maturation has been postulated to account for viral reinfections and failure of vaccines to protect against infection (28). Increasing SARS-CoV-2 antibody avidity over time has been demonstrated after COVID-19 infection (24,29,30). It is possible that the lower antibody avidity in AML patients seen in our study reflects incomplete antibody maturation in our immunocompromised patients, but longer-term longitudinal data are required to further investigate this hypothesis.

The desired result of vaccination is generation of effective neutralizing antibody activity. Vaccine effectiveness also depends on other aspects of the immune response, including amount of antibody produced, immunoglobulin subclasses, the affinity of the antibody for the target, and T-cell immune response. Khoury et al. have shown that SARS-CoV-2 antibody neutralization activity could be used to predict vaccine efficacy reported in phase 3 clinical trials (31). Our patients with AML had significantly less SARS-CoV-2 neutralizing antibody activity compared to healthy controls, and there was a trend for lower activity in MDS patients. Also, there was strong correlation between SARS-CoV-2 antibody level and neutralizing antibody activity (figure 5). Similarly strong correlation has also been reported by Malard et al. in patients with hematologic malignancies (32). Since quantification of neutralizing antibody activity may be difficult for clinical laboratories, quantitative SARS-CoV-2 antibody levels may be a good surrogate for neutralizing antibody activity. We did not find a correlation between antibody avidity and neutralizing activity (data not shown).

**Figure 5.**
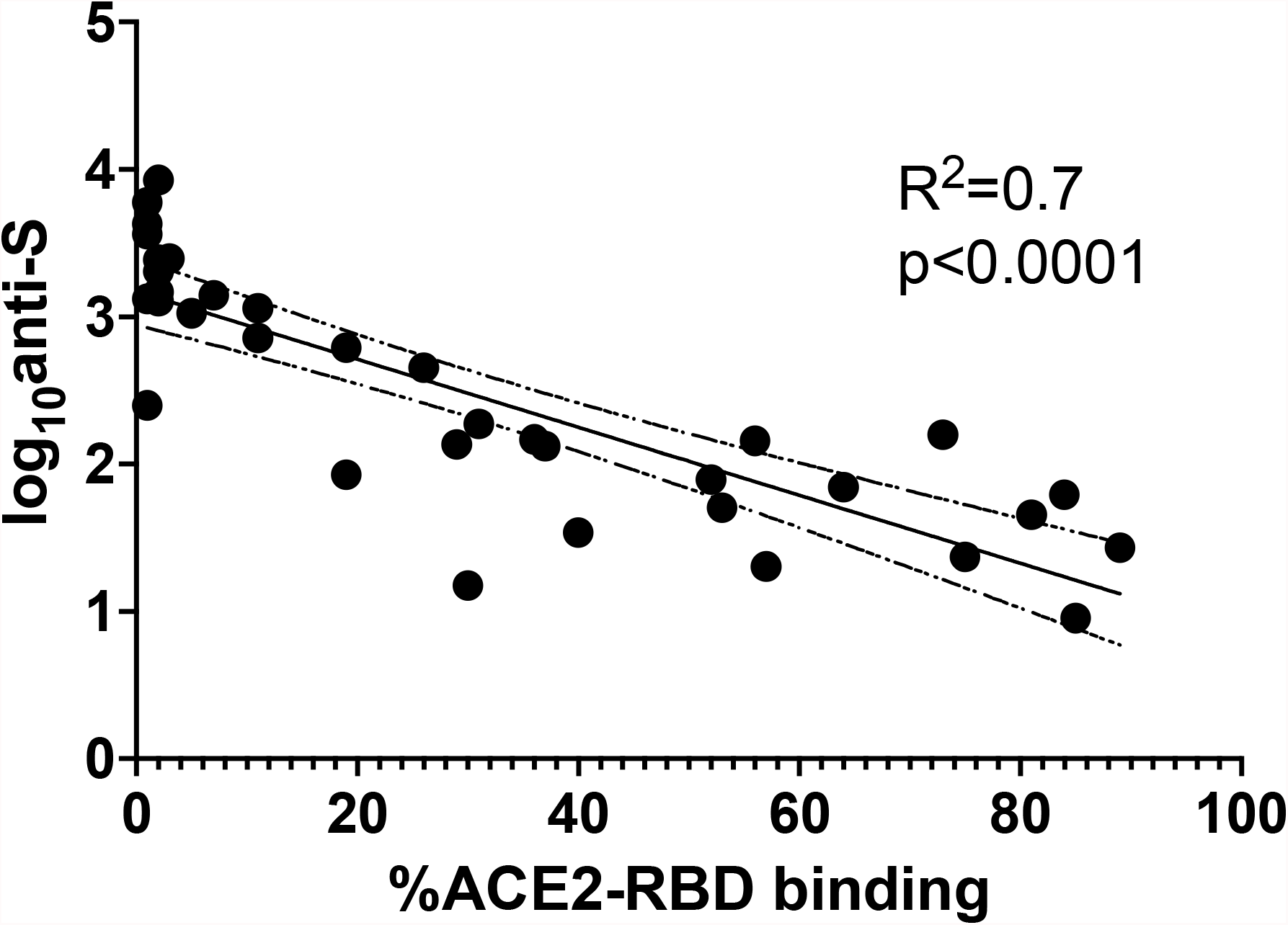
Anti-spike SARS-CoV-2 antibody titer correlates strongly with neutralizing antibody activity (R^2^=0.7, p<0.001) in the 35 patients with MDS and AML. Higher %ACE2-RBD binding represents lower neutralizing antibody activity. The dotted lines represent 95% confidence intervals.

In our study, 82% of pts with MDS or AML who received mRNA-1273 vaccine had an anti-spike SARS-CoV-2 antibody >100 U/ml, compared to only 50% of patients who received BNT162b2 vaccine. The median antibody level of patients receiving mRNA-1273 vaccine was more than 4-fold higher than for patients receiving BNT162b2 vaccine. Greenberger et al. found that a subset of non-Hodgkin’s lymphoma patients had higher seroconversion to the mRNA-1273 vaccine than to the BMT162b2 vaccine (15). In their study of patients with mostly CLL, lymphoma, and myeloma, Chung et al. noted that patients who received mRNA-1273 vaccine had higher SARS-CoV-2 anti-spike antibody titers compared to those who received BNT162b2 vaccine (23). Boyarski et al. found that solid organ transplant patients seroconverted more often to the first dose of mRNA-1273 vaccine compared to the first dose of BNT162b2 vaccine, but there was no difference in seroconversion rate after two doses (33). Recent reports in health care workers also demonstrated that vaccination with mRNA-1273 vaccine elicited higher antibody titers than vaccination with BNT162b2 vaccine (34,35). Zeng et al. found that a group of health care workers as well as a group of CLL and non-Hodgkin’s lymphoma patients who received mRNA-1273 vaccine had higher neutralizing antibody titers compared to the respective groups who received BNT162b2 vaccine (20). Recent real-world clinical data suggest that breakthrough COVID-19 infections (36) and Delta variant COVID-19 hospitalizations (37) may be more common in BNT162b2 vaccinated persons than mRNA-1273 vaccinated persons. Our results are consistent with these findings. Furthermore, we found that antibody avidity was significantly greater and SARS-CoV-2 neutralizing antibody activity was higher in our patients who received mRNA-1273 vaccine compared to BNT162b2 vaccine. Our results suggest that the immune response to mRNA-1273 vaccine may be more robust than the immune response to BNT162b2 vaccine in patients with AML and MDS. Whether this difference is due to vaccine mRNA concentration, vaccine timing, or something otherwise inherent to the vaccine remains to be determined. The clinical significance of this differential immune response is also unknown.

Vaccine efficacy for mRNA vaccines is reduced in immunocompromised patients compared to immunocompetent hosts. The BNT162b2 vaccine efficacy to prevent COVID-19 is 75% for immunocompromised hosts compared to 94% for immunocompetent hosts (38). Vaccine efficacy of the mRNA vaccines to prevent hospitalization from COVID-19 is 59% for immunocompromised hosts compared to 91% for immunocompetent hosts (39). The same study found that of previously vaccinated hospitalized patients with breakthrough COVID-19, 44% were immunocompromised (39). A multicenter study of breakthrough infections found that of hospitalized patients with COVID-19 who had received BNT162b2 vaccine, 40% were immunocompromised by chemotherapy, corticosteroids, anti-CD20 antibody, or solid organ transplant (40). The diminished immune response to COVID-19 vaccination in immunocompromised patients, as we have demonstrated with decreased antibody titers, antibody avidity, and neutralizing antibody activity, may contribute to reduced vaccine efficacy.

## Data Availability

All data produced in the present study are available upon reasonable request to the authors

## Acknowledgements

Dr. Paul Christos was partially supported by the following grant: Clinical and Translational Science Center at Weill Cornell Medical College (1-UL1-TR002384-01).

